# Evaluation of T Cell Immune Memory Response after Sinopharm (BBIBP-CorV), Gam-COVID-Vac (Sputnik V), and Heterologous Sputnik-V/mRNA-1273 (Moderna) COVID-19 Vaccination Schemes against Different SARS-CoV-2 Variants

**DOI:** 10.1101/2024.12.13.24318016

**Authors:** Matías J. Pereson, María Noel Badano, Florencia Sabbione, Irene Keitelman, Natalia Aloisi, Susana Fink, Lucas Amaya, Gabriel H. Garcia, Alfredo P. Martínez, Federico A. Di Lello, Patricia Baré

## Abstract

**Introduction:** The adaptive immune response plays a crucial role in resolution of viral infections; however, there is limited data on the T cell response to SARS-CoV-2 vaccines like Sinopharm (Sin), Sputnik-V (SpV), and especially for the heterologous combination Sputnik-V/mRNA-1273 (Spv/Mod) scheme. Furthermore, emerging variants may compromise the adaptive immune response established in the population.

**Objective:** To evaluate the T cell immune response induced by the Sin, SpV and SpV/Mod, against Wuhan, Gamma and Omicron viral variants.

**Methods:** Serum levels of IgG antibodies were assessed by CMIA (Abbott Diagnostics, Abbott Park, Illinois). T cell responses against Wuhan, Gamma, and Omicron were evaluated through an activation-induced marker assay. Sixty individuals, evenly distributed across the Sin, SpV, and SpV/Mod groups, were included.

**Results:** The median age of participants was 48 years (IQR: 34-56), with 58.3% (n=35) being female. Seventeen (28.3%) individuals had a prior confirmed COVID-19 infection. Spv/Mod scheme elicited significantly higher humoral responses (p<0.001). However, the three vaccination platforms showed consistent T cell responses across the different Wuhan, Gamma and Omicron stimuli tested. Particularly, all three schemes induced stronger CD8 responses against the Wuhan variant. In addition, more responders to Wuhan were observed in the Sin and Spv groups, while the Spv/Mod group showed a higher proportion of responders to Omicron.

**Conclusion:** This study provides new data on effective humoral and cellular immune responses against Wuhan, Gamma, and Omicron variants. Moreover, memory CD4 and CD8 T cell responses remain protective, and the heterologous scheme may elicit stronger T cell responses against emerging variants.

**Highlights:**

- Strong adaptive T cell responses were observed across all vaccination schemes and viral variants.
- Higher frequency of activated CD8 lymphocytes was detected compared to CD4 in response to SARS-CoV-2
- Sinopharm and Sputnik-V showed a response biased toward the Wuhan variant.
- Sputnik-V/Moderna recipients showed higher T cell responses to Omicron.
- The Sputnik-V/Moderna vaccination scheme induced significantly enhanced humoral immune responses.

**Graphical Abstract:** 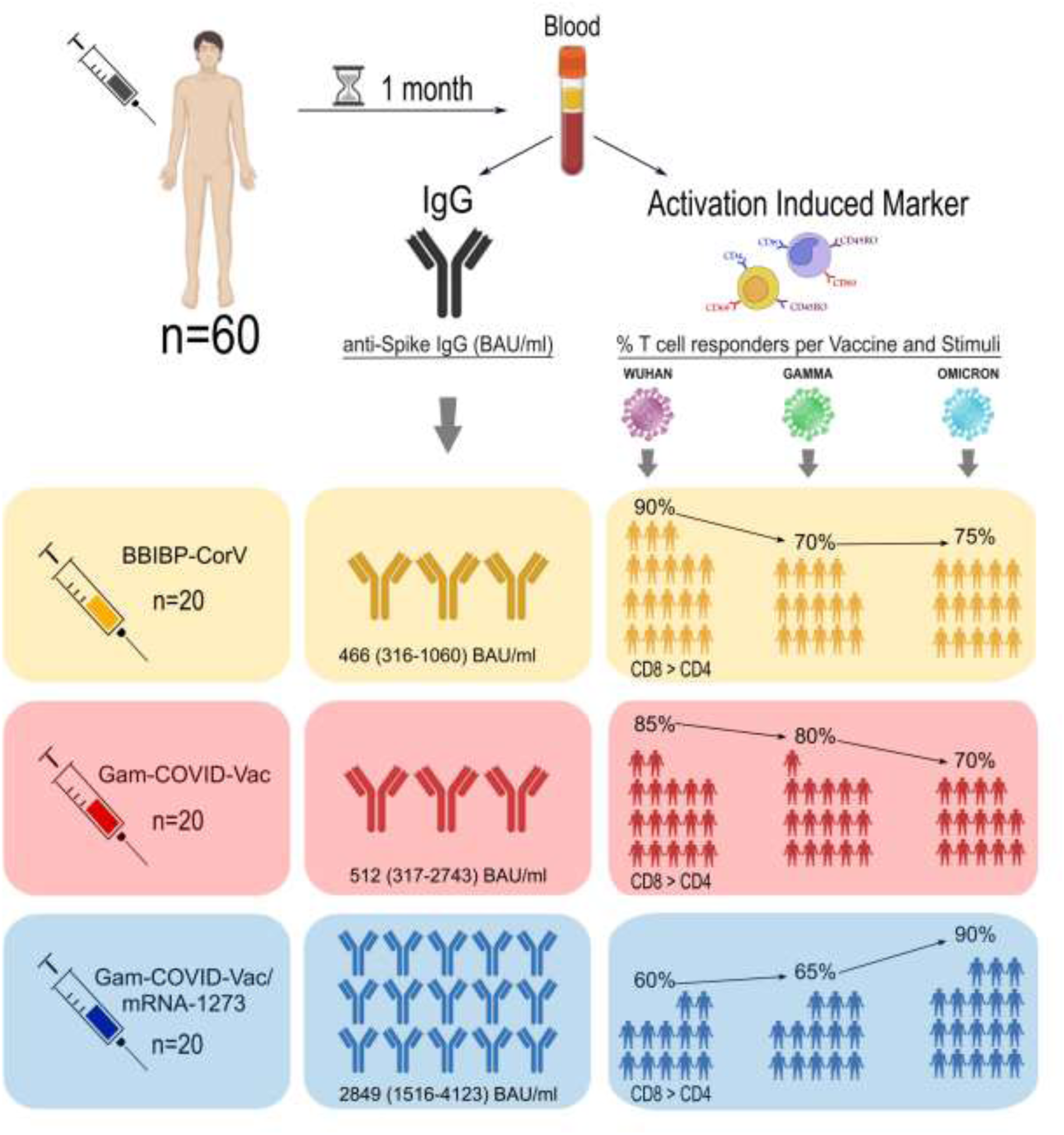

## Introduction

SARS-CoV-2-specific humoral and T cell responses are essential for protection against symptomatic and severe disease, with memory CD4 and CD8 T-lymphocytes providing long-term protection, even against emerging SARS-CoV-2 variants ^[1]^.

In Argentina, three major waves of SARS-CoV-2 infections occurred. The first began in late 2020, characterized by the predominance of ancestral Wuhan-like viral variants ^[2]^. The second in mid-2021, coinciding with the initial phases of vaccine campaign and the rise of Gamma variant ^[2]^. Common vaccination schemes in Argentina included BBIBP-CorV [Sinopharm (Sin)] and Gam-COVID-Vac [Sputnik-V (SpV)] homologous regimens. However, due to shortages of the second component of the Sputnik-V vaccine, the primary vaccination scheme was completed by administering a second dose of the mRNA-1273 [Moderna (Mod)] vaccine, mainly to elderly and other previously prioritized groups. This heterologous combination (Spv/Mod) was innovative both locally and globally ^[3]^. Shortly afterward, the third wave dominated by Omicron, irrupted in the population posing challenges to the recent immune responses induced by vaccination or prior infection ^[2]^.

Many studies have evaluated the T cell response to the Sinopharm ^[4–9]^ and Sputnik-V vaccines ^[5,10–13]^. However, research on the Sputnik-V/Moderna combination is limited ^[14]^ and even more scarce are comparative analysis of cellular responses generated by these three vaccination schemes. In addition, only two studies evaluated the cellular responses against variants of concern beyond the ancestral Wuhan virus, both focusing on Sin recipients against Omicron ^[6,7]^. Therefore, comparative studies involving multiple vaccine platforms and viral variants are needed. The aim of this study was to assess the CD4 and CD8 T cell memory immune responses elicited by the Sin, SpV, and heterologous SpV/Mod combination against the Wuhan, Gamma, and Omicron SARS-CoV-2 variants.

## Materials and Methods

### Study Population

This retrospective cohort study included healthcare workers of the Academia Nacional de Medicina and attendees of the Centro de Educación Médica e Investigaciones Clínicas “Norberto Quirno” (CEMIC). The cohort comprises individuals with or without a history of previous COVID-19, all of whom were vaccinated with two doses of the Sin, SpV or SpV/Mod schemes. Detailed information regarding vaccine dose administration and infection history (including date, laboratory confirmation, or epidemiological criteria) was obtained through surveys. Twenty pre-pandemic donors’ samples from 2018 and early 2019 were included as a negative control group (CG).

### Sample Collection and Processing

Peripheral blood samples were collected in tubes containing ethylenediaminetetraacetic acid (EDTA). Plasma was obtained via centrifugation to assess anti-SARS-CoV-2 IgG. Peripheral blood mononuclear cells (PBMCs) were isolated using Ficoll-Paque density gradient centrifugation, resuspended in RPMI 1640 medium with 57% inactivated fetal bovine serum (GIBCO-Thermo Fisher Scientific, CA, USA 92008), 10% dimethyl sulfoxide (DMSO), and cryopreserved in liquid nitrogen until use.

### Assessment of Cellular Immune Response against SARS-CoV-2

Flow cytometry was used to analyze CD69 activation marker expression on memory T cells (CD45RO, CD4, and CD8) after stimulation with SARS-CoV-2 receptor-binding domain (RBD) proteins from the ancestral Wuhan strain and the Gamma and Omicron variants.

Cryopreserved PBMCs were rapidly thawed and distributed into 96-well plates at a density of 5×10^5 cells per well in a final volume of 200μL of RPMI culture media supplemented with 10% fetal bovine serum, and 1% penicillin/streptomycin (Thermo Fisher Scientific, CA, USA), and incubated at 37°C and 5% CO2. After resting for 24h, the cells were stimulated with the corresponding SARS-CoV-2 proteins at a final concentration of 2.5 μg/mL. Unstimulated cells and cells stimulated with phytohemagglutinin (5 μg/mL, Sigma) were included as negative and positive controls, respectively. After 19 h, cells were harvested and stained with surface markers [anti-CD3 (BV-421), anti-CD4 (PE-Cy7), anti-CD8 (PerCP-Cy5.5), anti-CD45RO (FITC), and anti-CD69 (PE) antibodies, all from BD Pharmingen], fixed using the BD Cytofix/Cytoperm™ kit, and acquired on a FACScanto II cytometer (BD Pharmingen).

Data analysis was performed using FCS Express V5 software (De Novo Software, Los Angeles, CA, USA). Lymphocytes were gated based on their forward and side scatter (Forward Scatter vs. Side Scatter), and doublets were excluded. Subsequently, the frequencies of CD3+CD4+CD45RO+ and CD3+CD8+CD45RO+ T cells co-expressing the CD69 activation marker, corresponding to activated memory helper (CD4) and cytotoxic (CD8) T cells in response to the different tested stimuli, were analyzed. In addition, a threshold value was established by taking the median baseline value for each stimulus condition in the negative control group (pre-pandemic group) and adding one standard deviation. This threshold was used to determine whether individuals’ responses were positive or negative, as described in previous studies ^[15,16]^ yielding CD4 values of 0.231%, 0.202%, and 0.148%, and CD8 values of 0.5%, 0.195%, and 0.252% for the Wuhan, Gamma and Omicron stimuli, respectively. Values below the threshold (i.e., negative responses) in the vaccinated groups were excluded from the analysis.

### Assessment of Humoral anti-Spike IgG Antibody Concentration

Binding IgG antibodies targeting the SARS-CoV-2 spike (S) receptor-binding domain (RBD) were measured 3-7 weeks post-boost using the Abbott SARS-CoV-2 IgG II Quant chemiluminescent microparticle immunoassay (CMIA) on Architect i2000 SR and Alinity I analyzers. Results were standardized to World Health Organization (WHO) binding antibody units (BAU) using a conversion factor based on the WHO international standard NIBSC 20-136.

### Statistical Analysis

Data were analyzed using GraphPad Prism 10.3.1 (GraphPad Software, San Diego, CA, USA). The Wilcoxon signed-rank or Mann-Whitney U test was used to compare two variables, depending on whether the tests were related or independent. For comparisons among three or more groups, the Kruskal-Wallis or Friedman test was used, based on group dependency. The Shapiro-Wilk test was applied to check for normal distribution. P-values <0.05 were considered significant. Detailed statistical tests are provided in the figure legends.

### Ethical Considerations

The study was conducted in accordance with the Declaration of Helsinki, and all participants provided written informed consent. The experimental protocols were approved by the Biosafety Review Board, the Ethical Committee of the Academia Nacional de Medicina, Buenos Aires (T.I.N° 133335/21; 12/22/CEIANM) and by the Ethical Committee of the Faculty of Pharmacy and Biochemistry, University of Buenos Aires (EX-2021-06438339-UBA).

## Results

### Study Population

The cohort analyzed included 60 individuals who received a two-dose vaccination regimen in 2021: Sin (n=20), SpV (n=20), and SpV/Mod (n=20). The median age (IQR) was 48 (34-56) years. Among them, 35 (58.3%) were women and 17 (28.3%) individuals had a history of COVID-19. Age was significantly higher in the heterologous group (p=0.042). Table 1 shows participant characteristics by vaccination scheme.

**Table 1.** Characteristics of Study Populations according to Administered Vaccination Scheme (N=60).

| *Characteristics* | *Sinopharm*<br>*(n=20)* | *Sputnik*<br>*(n=20)* | *Sputnik/Moderna*<br>*(n=20)* | *Total*<br>*(N=60)* | *p* |
| --- | --- | --- | --- | --- | --- |
| *Age (years), Median (IQR)* | 45 (37-50) | 49 (32-60) | 56 (41-62) | 48 (34-56) | p=0.042 |
| *Female gender, (%)* | 15 (75) | 9 (45) | 11 (55) | 35 (58) | p=0.151 |
| *Prior COVID-19, (%)* | 7 (35) | 6 (30) | 4 (20) | 17 (28) | p=0.568 |
| *Time from last exposure*<br>*(days), Median (IQR)* | 24 (22-25) | 22 (20-31) | 21 (19-27) | 23 (21-28) | p=0.368 |

\* IQR = Interquartile Range (Q1-Q3)

### Cellular Immune Response against SARS-CoV-2 by Vaccination Scheme

The median frequencies for CD4 responses were 0.7% (IQR 0.4-1.17) for Sin, 0.97% (IQR 0.412-1.503) for SpV, and 0.68% (IQR 0.385-1.035) for SpV/Mod. For CD8, the median values were 1.34% (IQR 0.562-3.688) for Sin, 1.18% (IQR 0.685-2.115) for SpV, and 1.01% (IQR 0.575-1.953) for SpV/Mod. Activated T cells were significantly higher in the three vaccinated groups compared with the pre-pandemic control group (Figure 1). No differences were observed between schemes for CD4 or CD8 responses (p=0.512 and p=0.504, respectively). Higher percentages of activated CD8 T cells were observed compared to CD4 T cells in all three groups (Sin p=0.009, SpV p=0.039, and Spv/Mod p=0.030).

**Figure 1.**
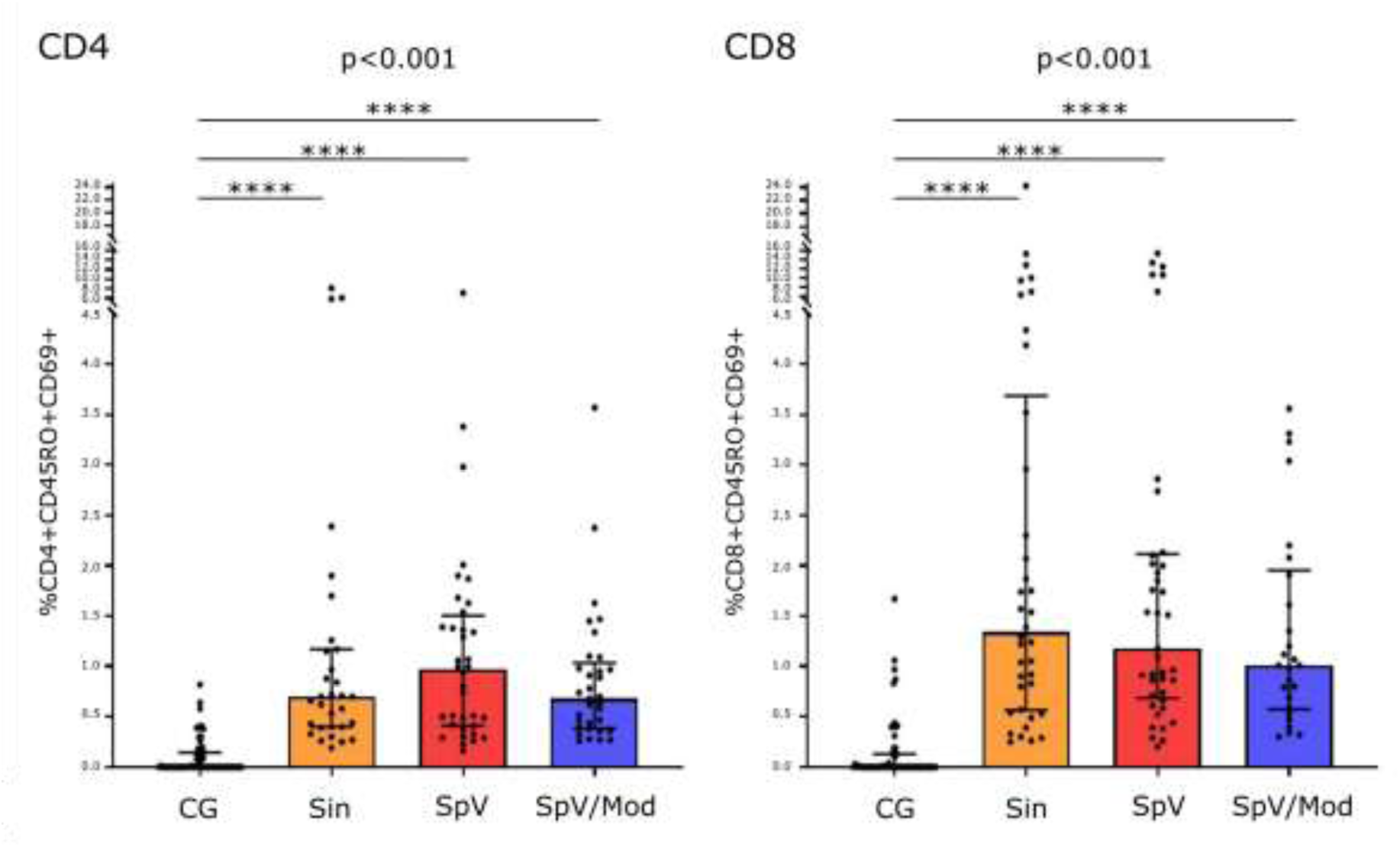
Graphical representation of SARS-CoV-2 specific CD4 and CD8 memory T-cellular responses from the primary vaccination schemes (Sinopharm, Sputnik, and Sputnik/Moderna) against all tested SARS-CoV-2 stimuli. **a.** Comparison of the frequencies (%) of CD4 lymphocytes from evaluated vaccination schemes against those obtained in the control group. **b.** Comparison of the frequencies (%) of CD8 lymphocytes from evaluated vaccination schemes against those obtained in the control group. Statistical analysis was performed using the non-parametric Kruskal-Wallis test. Pairwise comparisons were conducted using the Dunn’s multiple comparisons test. All comparisons between vaccinated groups yielded non-significant results. Values are expressed on a logarithmic scale for graphical purposes. The number of positives responses for CD4 were n=31 for Sin, n=36 for SpV, and n=33 for SpV/Mod. For CD8, the positives responses were n=38 for Sin, n=41 for SpV, and n=26 for SpV/Mod. CG=control group.

### Activated CD4 and CD8 T Cell Population Frequencies according to Stimuli and Vaccination Platforms

Subjects vaccinated with Sin showed higher percentages of CD4 lymphocytes for Wuhan and Gamma compared to Omicron (p=0.028). In the SpV group, higher percentages of CD4 lymphocytes were observed in response to the Wuhan compared to Gamma and Omicron (p=0.039). For the SpV/Mod scheme, no significant differences were observed between different stimuli for the CD4 response (p=0.374) (Table 2). For the CD8 compartment, higher percentages of activated T cells against Wuhan were observed across all three groups, although the difference for Spv/Mod did not reach statistical significance (Sin: p<0.001, SpV; p<0.001, and SpV/Mod; p=0.075). Moreover, no significant differences were observed among the vaccination schemes in the percentages of activated CD4 or CD8 lymphocytes in response to the same stimulus (Table 2).

**Table 2.** Frequency of activated CD4 and CD8 memory T lymphocytes from different vaccination schemes in response to different SARS-CoV-2 stimuli.

| CD4 |  | Sinopharm |  | Sputnik-V |  | Sputnik-V/Moderna |  | p (inter scheme) |
| --- | --- | --- | --- | --- | --- | --- | --- | --- |
|  |  | Median | IQR | Median | IQR | Median | IQR |  |
|  | Wuhan | 0.720 | 0.440-1.90 | 1.315 | 0.785-1.763 | 0.920 | 0.555-1.373 | 0.241 |
|  | Gamma | 0.775 | 0.590-1.518 | 0.455 | 0.298-1.383 | 0.710 | 0.355-1.419 | 0.294 |
|  | Omicron | 0.345 | 0.255-0.625 | 0.745 | 0.300-1.288 | 0.470 | 0.375-0.845 | 0.238 |
| p (intra scheme) |  | 0.028 |  | 0.039 |  | 0.374 |  |  |

| CD8 |  | Sinopharm |  | Sputnik-V |  | Sputnik-V/Moderna |  | p (inter scheme) |
| --- | --- | --- | --- | --- | --- | --- | --- | --- |
|  |  | Median | IQR | Median | IQR | Median | IQR |  |
|  | Wuhan | 4.340 | 2.070-10.01 | 2.860 | 1.850-10.72 | 2.200 | 1.305-3.300 | 0.381 |
|  | Gamma | 0.570 | 0.360-1.395 | 0.740 | 0.485-1.345 | 0.775 | 0.385-1.178 | 0.924 |
|  | Omicron | 0.970 | 0.470-1.593 | 0.910 | 0.480-1.730 | 1.01 | 0.790-1.610 | 0.780 |
| p (intra scheme) |  | <0.001 |  | <0.001 |  | 0.075 |  |  |

\* IQR = Interquartile Range (Q1-Q3)

### Number of Individuals who Responded to Different Stimuli

A positive responder was defined as a subject with a response exceeding the established threshold in any of the CD4, CD8, or both CD4 and CD8 compartments in response to the stimuli (Wuhan, Gamma or Omicron). Overall, 19 (95%) individuals vaccinated with Sin, 18 (90%) with SpV, and 19 (95%) with SpV/Mod showed at least one positive response to any stimuli.

When evaluating each viral variant across vaccine platforms, and considering that the heterologous combination Spv/Mod was innovative with limited reports on its efficacy against different variants, we compared the heterologous to the homologous scheme. We found that the proportion of responders to the Wuhan variant in Spv/Mod was significantly lower than in the Sin and Spv groups (p=0.022). In contrast, the proportion of responders to the Gamma variant showed no significant differences among the groups (p=0.545). Interestingly, for Omicron variant, the SpV/Mod scheme demonstrated a trend toward a higher proportion of responders compared to the Sin and Spv groups (p=0.083)

Taking into account the response to the three different stimuli for each vaccine platform, the Sin and Spv groups (orange and red line in Figure 2) showed a trend toward higher number of responders against Wuhan compared to Gamma and Omicron (p=0.156 and p=0.246 respectively). The SpV/Mod group (blue line in Figure 2) exhibited a tendency toward a higher number of responders to the Omicron variant compared to the Wuhan and Gamma variants (p=0.059) (Figure 2).

**Figure 2.**
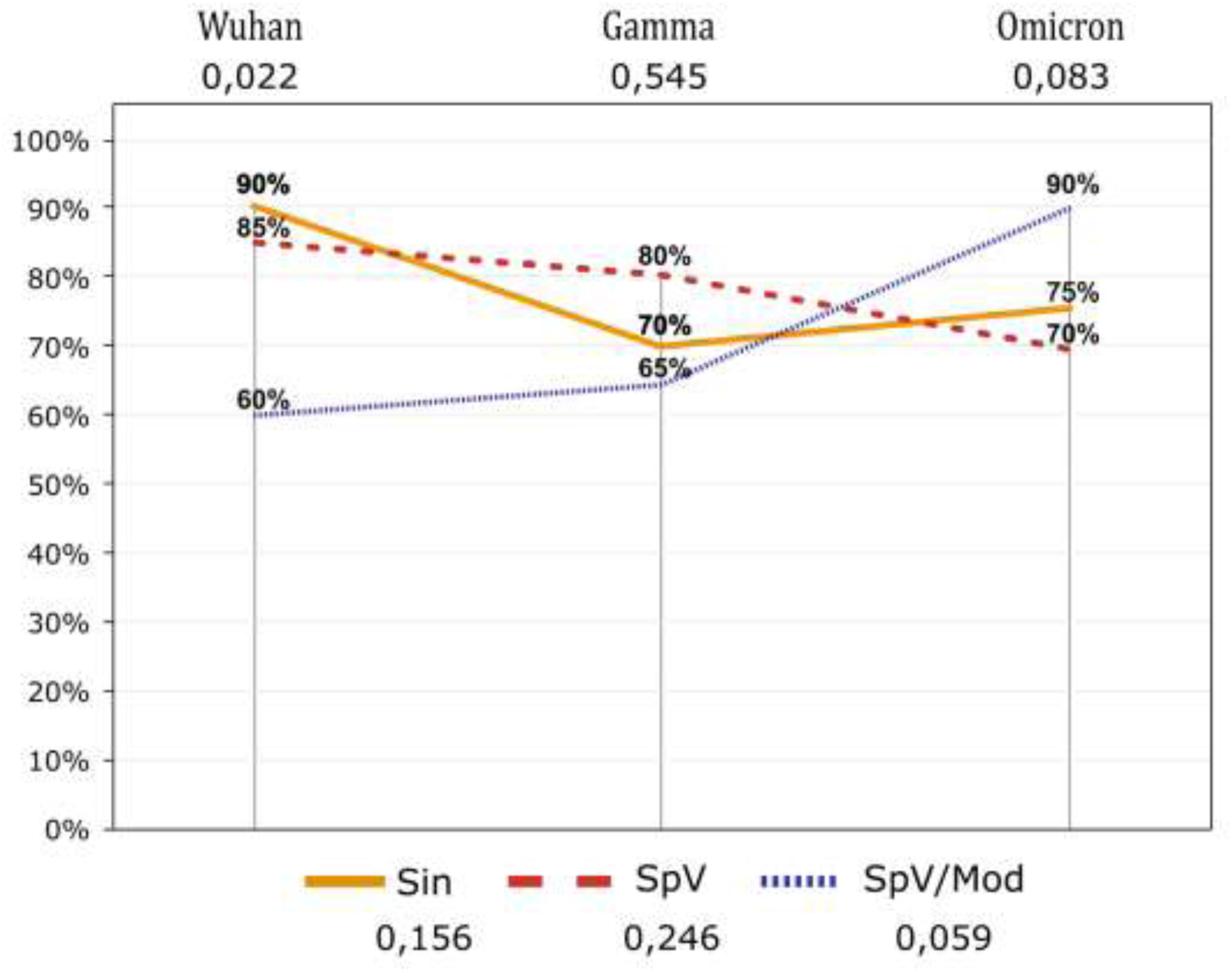
Graphical representation of the number of responders by vaccination schemes and tested stimuli. Sin = Sinopharm, SpV = Sputnik-V, SpV/Mod = Sputnik-V/Moderna. W = Wuhan, G = Gamma, and O = Omicron.

### Assessment of Humoral Anti-Spike IgG Antibody Concentration

Median IgG titers were significantly higher in the SpV/Mod group (2849 BAU/mL, IQR 1516-4123) compared to SpV (512 BAU/mL, IQR 317-2743, p=0.001) and Sin (466 BAU/mL, IQR 316-1060, p<0.001). No significant differences were found in the humoral response elicited by Sin and Spv (p=0.808). Individuals with a history of COVID-19 infection developed higher antibody titers compared to vaccinated individuals who had not been infected, although the difference was not statistically significant for the Sin group (Table 3).

**Table 3.** Humoral IgG responses in SARS-CoV-2 naïve and previously infected individuals, categorized by vaccination scheme.

|  | Naïves | Prior COVID-19 | p |
| --- | --- | --- | --- |
| Sinopharm* | 398 (290-536) | 1441 (313-1930) | 0.081 |
| Sputnik-V* | 395 (282-525) | 4028 (2743-5063) | <0.001 |
| Moderna* | 2765 (1415-4269) | 4747 (3677-5636) | 0.021 |

\* IQR = Interquartile Range (Q1-Q3)

## Discussion

The present study shows that while both the Sin and SpV vaccines induced strong T cell responses against the Wuhan variant, SpV/Mod may generate a stronger T cell response to Omicron. Additionally, the heterologous scheme induced higher humoral responses. These results provide new and original data underscoring the significance of this study.

When evaluating the cellular immune response, we observed that over 90% of participants exhibited a positive T cell response across the three schemes evaluated. Specifically, 90% of individuals vaccinated with Sin showed T cell responses against Wuhan. This result is concordant with previous studies that detected responses in the range of 28.6% to 100% against the ancestral SARS-CoV-2 strain ^[4,5,7,14]^. Additionally, two studies also evaluated the T cell response of Sin platform against Omicron. Li et al. reported Omicron-specific T cell responses in 26.7% of the 15 individuals evaluated ^[6]^, while Lim et al. observed it in 72% of the 18 subjects evaluated ^[7]^. Similarly, in our study, we observed a strong T cell response against Omicron in the Sin group, with 75% of responders. Furthermore, the Sin group also displayed a strong response against Gamma (70%), comparable to the response against Omicron, but slightly weaker than the response against Wuhan. To our knowledge, no studies have evaluated T cell responses of Sin-vaccinated individuals against the Gamma variant for comparison.

In our study, 85% of the individuals vaccinated with SpV showed a T cell response against Wuhan, consistent with previous publications reporting responses between 40% and 100% ^[4,9,10,12–14]^. No reports have yet evaluated the SpV cellular response against emerging viral variants. Here, we observed strong T cell memory responses of the SpV group against Gamma (80%) and Omicron (70%). Both, the Sin and SpV vaccination schemes, yielded similar results in terms of the number of responders and the percentages of activated CD4 and CD8 T-lymphocytes against these variants, with stronger responses against Wuhan and a trend of decreasing response to newer variants. This phenomenon may be explained by the reduced T cell recognition due to mutations within the RBD protein ^[17,18]^.

Only one study has evaluated T cell responses of the SpV/Mod heterologous scheme, reporting a 44% responder rate in 18 individuals evaluated against Wuhan ^[14]^. In our study, a lower percentage of T cell responders was observed against the Wuhan and Gamma variants for the heterologous group compared to the number of responders seen in the Sin and SpV groups. In this line, Nuñez et al observed that T cell responses predominated in the Sin group, while the SpV/Mod combination enhanced B cell and humoral responses ^[14]^. Notably, almost all SpV/Mod recipients responded to Omicron (90%), a finding that may warrant further investigation. In this regard, a recent report found that a heterologous vaccination regimen combining Janssen’s viral vector platform and Pfizer’s mRNA platform generated a more diverse lymphocytes clonal repertoire compared to a homologous vector platform, potentially providing greater protection against emerging SARS-CoV-2 variants ^[19]^. Age-related factors, such as previous infections with other *coronaviruses*, may influence cross-reactive T cell responses to different SARS-CoV-2 variants. This limitation cannot be addressed as vaccination distribution was based on risk priorities set by the Ministry of Health.

Furthermore, all three-vaccination schemes showed higher frequencies of activated CD8 memory T cells compared to activated CD4 T cells. Previous studies show mixed results, with some reporting a CD4 bias ^[7,10]^, others higher CD8 responses ^[8,20]^, and some finding no differences ^[9]^. These discrepancies stem from variations in T cell evaluation methods.

Evaluating T cell responses in vaccinated individuals is essential, particularly in order to assess their capacity to respond to emerging viral variants. Considering Argentina’s epidemiological history, our findings suggest that these vaccination schemes have likely established strong immunity against different SARS-CoV-2 variants circulating in our country, contributing to population protection against severe disease.

In addition, the results on humoral response showed that all subjects, regardless of their vaccination scheme, developed detectable IgG antibodies against SARS-CoV-2, concordant with previous studies reporting seroconversion in nearly all individuals vaccinated with these platforms ^[4,9,11,14]^. Additionally, higher antibody titers were observed in individuals with prior infection and those who received the heterologous vaccine combinations. The enhancement of the humoral response after vaccination in individuals previously infected with SARS-CoV-2 is well documented ^[11,12]^. Moreover, the stronger humoral response observed in the SpV/Mod group reflects the effectiveness of the mRNA platform in generating a more robust humoral immune response ^[4,9,14,21–23]^.

In conclusion, the Sinopharm, Sputnik V, and Sputnik V/Moderna vaccination schemes effectively promote the development of both humoral and cellular immune responses. Additionally, the comparative study revealed that memory CD4 and CD8 T cell responses remain robust against Wuhan, Gamma and Omicron variants and suggest that the heterologous combination may offer greater effectiveness against emerging variants. In this regard, the strategy of combining vaccines has shown promise in the vaccination efforts for other viral infections and warrants further consideration in future immunization plans ^[24,25]^. Moreover, evaluating the T cell immune response is a valuable tool for achieving a more comprehensive understanding of vaccine efficacy. Finally, comparing different vaccine platforms against various variants can inform decision-making in shaping future national vaccination strategies.

## Data Availability

All data produced in the present work are contained in the manuscript.

## Acknowledgments

MJP, FS, IK, SF, MNB, FAD and PB are members of the National Research Council (CONICET) Research Career Program.

The authors thank all subjects enrolled in this study for their participation and collaboration. We are grateful to the COVIDAR group, Dr Andrea Gamarnik and Laboratorios Pablo Cassará for providing the purified proteins used for cell stimulation. Some aspects of this work could not have been fulfilled without the generous contribution of Dr Fernanda Palacios, Margot Peña and Diana Soto, and the technical support of Becton Dickinson Argentina S.R.L.

## Declaration of Author Contributions

MJP, FAD and PB conceived the study and designed the study protocol; MJP, MNB, NA, FS, IK, LA and AM carried out the clinical assessment; MJP, FAD and PB made the analysis and interpretation of the database. MJP, FAD and PB drafted the manuscript. MNB, SF, GG and AM critically revised the manuscript for intellectual content. All authors read and approved the final manuscript. MJP, FAD and PB are guarantors of the paper.

